# Genetic Variation in Blood Pressure and Lifetime Risk of Peripheral Artery Disease: A Mendelian Randomization Study

**DOI:** 10.1101/2020.08.23.20180240

**Authors:** Michael G. Levin, Derek Klarin, Venexia M. Walker, Dipender Gill, Julie Lynch, Kyung M. Lee, Themistocles L. Assimes, Pradeep Natarajan, Adriana M. Hung, Todd Edwards, Daniel J. Rader, J. Michael Gaziano, Neil M. Davies, Philip S. Tsao, Kyong-Mi Chang, Benjamin F. Voight, Scott M. Damrauer, On behalf of the VA Million Veteran Program

## Abstract

**Aims:** We aimed to estimate the effect of blood pressure and blood pressure lowering medications (via genetic proxies) on peripheral artery disease.

**Methods and Results:** GWAS summary statistics were obtained for BP (International Consortium for Blood Pressure + UK Biobank GWAS; N = up to 757,601 individuals), peripheral artery disease (PAD; VA Million Veteran Program; N = 24,009 cases, 150,983 controls), and coronary artery disease (CAD; CARDIoGRAMplusC4D 1000 Genomes; N = 60,801 cases, 123,504 controls). Genetic correlations between systolic BP (SBP), diastolic BP (DBP), pulse pressure (PP) and CAD and PAD were estimated using LD score regression. The strongest correlation was between SBP and CAD (r_g_ = 0.36; p = 3.9 × 10^−18^). Causal effects were estimated by two-sample MR using a range of pleiotropy-robust methods. Increased SBP, DBP, and PP increased risk of both PAD (SBP OR 1.25 [1.19–1.31] per 10mmHg increase, p = 3 × 10^−18^; DBP OR 1.27 [1.17–1.39], p = 4 × 10^−8^; PP OR 1.51 [1.38–1.64], p = 1 × 10^−20^) and CAD (SBP OR 1.37 [1.29–1.45], p = 2 × 10^−24^; DBP OR 1.6 [1.45–1.76], p = 7 × 10^−22^; PP OR 1.56 [1.4–1.75], p = 1 × 10^−15^). The effects of SBP and DBP were greater for CAD than PAD (p_diff_ = 0.024 for SBP, p_diff_ = 4.9 × 10^−4^ for DBP). Increased liability to PAD increased PP (beta = 1.04 [0.62–1.45] mmHg per 1 unit increase in log-odds in liability to PAD, p = 1 × 10^−6^). MR was also used to estimate the effect of BP lowering through different classes of antihypertensive medications using genetic instruments containing BP-trait associated variants located within genes encoding protein targets of each medication. SBP lowering via calcium channel blocker-associated variants was protective of CAD (OR 0.38 per 10mmHg decrease in SBP; 95% CI 0.19–0.77; p = 0.007).

**Conclusions:** Higher BP is likely to cause both PAD and CAD but may have a larger effect on CAD risk. BP-lowering through calcium-channel blockers (as proxied by genetic variants) decreased risk of CAD.

## INTRODUCTION

Peripheral artery disease (PAD) is a common manifestation of atherosclerotic cardiovascular disease (ASCVD), estimated to affect more than 12 million individuals in the United States, and more than 120 million individuals worldwide (1,2). PAD shares a number of risk factors with other forms ASCVD like coronary artery disease (CAD) and ischemic stroke (3). These risk factors include smoking, diabetes, hypertension, hyperlipidemia, and obesity (2–4). Observational studies have identified hypertension as one of the strongest risk factors for incident and prevalent PAD (5–11), although these studies may be limited by residual environmental confounding or reverse-causality. While randomized controlled trials of antihypertensive medications have demonstrated broad protection from coronary artery disease and death from cardiovascular causes, whether lower blood pressure reduces risk of PAD specifically has not been reliably established. Similarly, the relative effect of blood pressure on PAD and CAD has not been fully investigated.

Recent genome-wide association studies (GWAS) of PAD, CAD, and blood pressure including more than 700,000 individuals have identified hundreds of genetic variants associated with these traits (12,13). The Mendelian randomization (MR) framework (under certain assumptions) can leverage this genetic variation (which is randomly assorted during meiosis, mimicking a randomized trial), to provide unconfounded causal estimates of the relationship between traits (14). MR assumes that genetic variants are likely to be independent of many confounders of the exposure-outcome relationship. This assumption is plausible because genetic variants are randomly inherited by offspring from parents during meiosis and conception, analogous to treatment allocation in a randomized trial. Because large randomized trials evaluating the relationship between treatment of hypertension and PAD outcomes may be unfeasible, other study designs are needed to fill this evidence gap. Here, we leverage population-scale genetic variation within the Mendelian randomization framework to 1) establish the relationship between blood pressure and risk of PAD, 2) quantify differences in the effect of blood pressure on CAD and PAD, and 3) estimate the effect of blood pressure lowering (using genetic proxies of antihypertensive medications) on PAD risk.

## METHODS

### Study Exposures

The 2018 Evangelou et al. International Consortium for Blood Pressure + UK Biobank GWAS, which included measurements of systolic blood pressure, diastolic blood pressure, and pulse pressure in up to 757,601 individuals was used to identify genetic variants associated with the primary blood pressure phenotypes (15). This study included up to 299,024 European participants from 77 independent studies genotyped with various arrays and imputed to either the 1000 Genomes Reference Panel or the HRC panels, and 458,577 participants from the UK Biobank imputed to UK10K + 1000 Genomes reference panel. Measurement of blood pressure varied among cohorts, and study-specific details are presented in the supplemental material of Evangelou et al., 2018. Summary statistics for the blood pressure genome wide association study are publicly available, and may be downloaded from the NHLBI GRASP catalog (https://grasp.nhlbi.nih.gov/FullResults.aspx).

### Study Outcomes

The 2019 Million Veteran Program genome wide association study of Peripheral Artery Disease by Klarin et al. identified 24,009 cases and 150,983 controls of European Ancestry (12). This study defined cases and controls based on electronic health record phenotyping within the Veterans Affairs (VA) Healthcare System and was validated against ankle brachial index measurement and manual chart review. The current analysis focused on participants of European ancestry. MVP PAD genome wide association study summary statistics are available on dbGAP (Accession phs001672.v2.p1).

Genome-wide association study summary statistics for coronary artery disease were obtained from the Nikpay et al. 2016 CARDIoGRAMplusC4D 1000 Genomes-based GWAS. This study was a meta-analysis including 60,801 CAD cases and 123,504 controls, with genotypes imputed using the 1000 Genomes phase 1 version 3 reference. Summary statistics were downloaded from http://www.cardiogramplusc4d.org/data-downloads/.

### Genetic Correlation

LD-score regression was used to estimate the genetic correlation between blood pressure traits, and between blood pressure traits and ASCVD traits (https://github.com/bulik/ldsc) (16,17). GWAS summary statistics were obtained for systolic blood pressure, diastolic blood pressure, pulse pressure, peripheral artery disease, and coronary artery disease (12,13,15). Summary statistics were filtered to HapMap3 SNPs, and SNP heritability and genetic correlations were estimated using pre-computed 1000 Genomes European LD-scores.

### Mendelian Randomization

Two-sample Mendelian randomization analyses were performed in R using the *TwoSampleMR* package (https://github.com/MRCIEU/TwoSampleMR) (18). Genetic instruments for SBP, DBP, and PP were constructed using linkage-disequilibrium independent (r^2^ < 0.001, distance = 10,000kb; 1000 Genomes European reference panel), genome-wide significant (p < 5×10^−8^) variants identified using GWAS summary statistics for each trait **(Supplemental Table 3)**. For bi-directional MR analysis, additional instruments were constructed for CAD and PAD using the same procedure **(Supplemental Table 6)**. For each variant included in the genetic instruments, proportion of variance explained was calculated using the formula 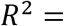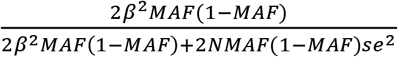(where MAF represents the effect allele-frequency, beta represents the effect estimate of the genetic variant in the exposure GWAS, se represents the standard error of effect size for the genetic variant, and N represents the sample size)(19). F-statistics were calculated for each variant using the formula 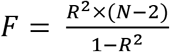 (where R^2^ represents the variance in exposure explained by the genetic variant, and N represents the number of individuals in the exposure GWAS) to assess the strength of the selected instruments **(Supplemental Table 4)**(20). For the primary analysis, power calculation was performed to identify the minimum-detectable effect size maintaining 80% power with two-sided alpha of 0.05. The primary MR analyses used inverse-variance weighting with random effects. The MR-Egger intercept test was used to evaluate for evidence of horizontal pleiotropy. Leave-one-out, single-SNP, and funnel-plot diagnostic MR analyses were performed. Sensitivity analyses were performed using MR methods that make different assumptions about the presence of pleiotropy (weighted median, penalized weighted median, and weighted mode) (21).

Multivariable MR (MVMR) was used in additional sensitivity analyses to jointly estimate the direct effects of multiple blood pressure traits, using genetic instruments including genomically independent (r^2^ < 0.001, distance = 10,000kb) variants that were associated at genome-wide significance (p < 5×10^−8^) with any exposure, weighted by the effect of each SNP on each exposure **(Supplemental Table 8)**(22). Effect estimates were scaled to correspond to a 10mmHg change in blood pressure.

### Antihypertensive Drug MR

Two MR analyses were performed to estimate the effect of 10mmHg lower of blood pressure by antihypertensive drugs. In the first analysis, genetic instruments consisting of variants within genes encoding protein targets of ACE-inhibitors, beta-blockers, and calcium-channel blockers were obtained from Gill et. al., with each variant weighted by its effect on systolic blood pressure **(Supplemental Table 5)**(23). In a sensitivity analysis, genetic instruments were constructed that mimic the action of 12 antihypertensive medication classes. In contrast to the first method which selected genetic variants based on their proximity to genes encoding protein targets, this method prioritized variants representing expression quantitative trait loci (eQTL) for genes encoding protein targets of antihypertensive medications that were demonstrated to effect systolic blood pressure (24). For both methods, inverse-variance weighted and weighted-median MR was performed, with MR-Egger intercept test used to assess for horizontal pleiotropy. For instruments with only 1 variant, Wald-ratio MR was performed.

### Statistical Analysis

All statistical analyses were performed using R version 3.6.2 (R Foundation for Statistical Computing).

## RESULTS

### Genetic Correlation

Cross-trait LD-score regression was used to estimate the genetic correlation between blood pressure traits. All blood pressure traits (SBP, DBP, PP) were strongly, positively correlated **(Supplemental Figure 1; Supplemental Table 1)**. The strongest correlation among blood pressure traits was between systolic blood pressure and pulse pressure (rg = 0.85; p < 1.0 × 10^−300^).

Genetic correlation between blood pressure traits and ASCVD traits (CAD and PAD) was then assessed. All BP traits were strongly, positively correlated with ASCVD traits **(Supplemental Figure 2; Supplemental Table 2)**. The strongest correlation among BP and ASCVD traits was between systolic blood pressure and CAD (rg = 0.36; p = 3.9 × 10^−18^).

### Effects of Blood Pressure on ASCVD: Mendelian Randomization

To determine whether the genetic correlations between BP and ASCVD traits would be consistent with causal effects, we performed two-sample mendelian randomization using genome-wide association study summary statistics. Genetic instruments for blood pressure contained between 342 and 410 independent genetic variants, explaining between 3.5% and 4.3% of the variability in blood pressure, with F-statistics ranging from 29.6 to 670.6 (consistent with low risk of weak-instrument bias) **(Supplemental Tables 3–4)**. The primary analysis maintained power to detect a 7% – 11% increase in risk of ASCVD per 10mmHg increase in blood pressure **(Supplemental Table 4)**. In inverse-variance weighted analyses, each 10mmHg increase in SBP increased risk of both PAD (OR 1.25; 95% CI 1.19–1.31; p = 3 × 10^−18^) and CAD (OR 1.37; 95% CI 1.29–1.45; p = 2 × 10^−24^), though the effect was stronger for CAD than PAD (pdifference = 0.024) **(Figure 1, Supplemental Table 5)**. Each 10mmHg increase in DBP increased risk of both PAD (OR 1.27; 95% CI 1.17–1.39; p = 4 × 10^−8^) and CAD (OR 1.6; 95% CI 1.45–1.76; p = 7 × 10^−22^), with a stronger effect for CAD than PAD (pdifference = 4.9 × 10^−4^) **(Figure 1, Supplemental Table 5)**. Each 10mmHg increase in PP increased risk of both PAD (OR 1.51; 95% CI 1.38–1.64; p = 1 × 10^−20^) and CAD (OR 1.56; 95% CI 1.4–1.75; p = 1 × 10^−15^), with similar effects on CAD and PAD (pdifference = 0.60) **(Figure 1, Supplemental Table 5)**. MR-Egger bias intercept term was p > 0.05 for all trait-outcome pairs except PP-PAD (p = 0.012) **(Supplemental Table 5)**. Results remained robust in sensitivity analyses using MR methods that make different assumptions about the presence of pleiotropy **(Supplemental Table 5)**.

**Figure 1:**
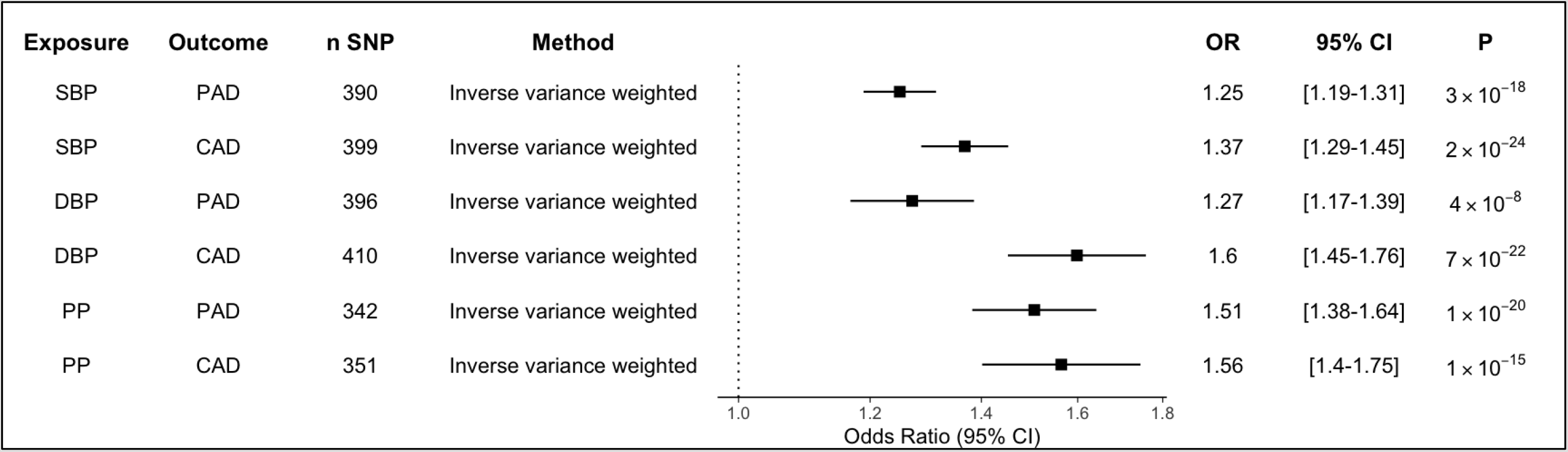
Effect of Blood Pressure Traits on ASCVD Outcomes. In inverse variance weighted Mendelian randomization analyses, elevations in each blood pressure trait increased risk of both coronary and peripheral artery disease. Results scaled to reflect odds of outcome per 10mmHg increase in blood pressure. n SNP = number of single nucleotide polymorphisms in the exposure instrument; OR = odds ratio; 95% CI = 95% confidence interval; P = pvalue SBP = Systolic blood pressure; DBP = Diastolic blood pressure; PP = Pulse pressure; PAD = Peripheral artery disease; CAD = coronary artery disease.

### Effects of Liability to ASCVD on Blood Pressure: Mendelian Randomization

Because stiffening of peripheral vessels in the setting of peripheral artery disease may affect blood pressure, raising the possibility of reverse-causation in assessment of the relationship between blood pressure and PAD, bi-directional MR analysis was performed. Genetic instruments for PAD and CAD were selected and used to estimate the effect of liability to ASCVD on blood pressure traits **(Figure 2; Supplemental Tables 6–7)**. In inverse-variance weighted analysis, liability to PAD increased PP (beta = 0.97 mmHg per 1 log-odds increase in risk of PAD; 95% CI 0.4–1.5; p = 8 × 10^−4^), and CAD increased PP (beta = 0.542 per 1 log-odds increase in risk of CAD; 95% CI 0.006–1.1; p = 0.05. Neither PAD nor CAD affected SBP or DBP. The MR-Egger bias intercept term had p > 0.05 for all analyses, indicating no positive evidence for bias. In sensitivity analysis applying MR methods making different assumptions about the presence of pleiotropy, there was weak evidence that both PAD and CAD increase SBP and PP while decreasing DBP **(Supplemental Table 7)**.

**Figure 2:**
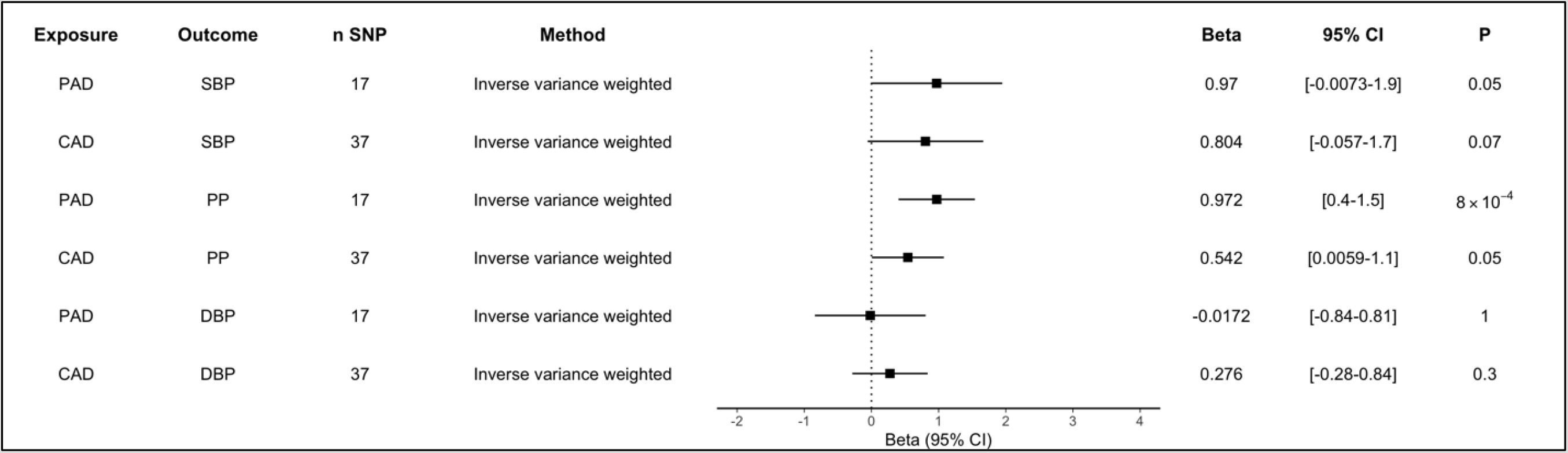
Effect of ASCVD Traits on Blood Pressure. In inverse variance weighted Mendelian randomization analyses, both peripheral artery disease and coronary artery disease increased pulse pressure. Results reflect increase in blood pressure (mmHg per 1 log-odds unit increase in risk of the exposure). n SNP = number of single nucleotide polymorphisms in the exposure instrument; OR = odds ratio; 95% CI = 95% confidence interval; P = p-value SBP = Systolic blood pressure; DBP = Diastolic blood pressure; PP = Pulse pressure; PAD = Peripheral artery disease; CAD = coronary artery disease.

### Multivariable Mendelian Randomization

Because blood pressure traits are highly correlated and unlikely to affect cardiovascular outcomes in isolation, we performed multivariable MR to jointly estimate the direct effects of each blood pressure trait on ASCVD outcomes **(Supplemental Table 8)**. Each 10mmHg increase in SBP increased risk of both PAD (direct OR 1.67; 95% CI 1.19–2.34; p = 0.003) and CAD (direct OR 1.44; 95% CI 1.02–2.03; p = 0.04). After accounting for the effect of SBP, we did not observe residual evidence of a significant direct effect of DBP on PAD or CAD.

### Antihypertensive Drug Mendelian Randomization

Two Mendelian randomization analyses were performed to estimate the effect of blood pressure lowering on ASCVD outcomes. The first analysis focused on the effect of SBP lowering using genetic variants located within genes encoding protein targets of common classes of blood pressure lowering medications (ACE-inhibitors, beta-blockers, and calcium channel blockers) **(Figure 3; Supplemental Tables 10–11)**. This analysis identified protective effects of beta-blocker-associated variants on CAD (OR 0.59 per 10mmHg decrease in SBP; 95% CI 0.44- 0.78; p = 3 × 10^−4^), and protective effects of calcium channel blocker associated variants on CAD (OR 0.69 per 10mmHg decrease in SBP; 95% CI 0.59–0.80; p = 2 × 10^−6^). No effect of blood pressure lowering via genetic proxies of these medication classes on risk of PAD was detected. Results were consistent in weighted-median MR sensitivity analyses **(Supplemental Table 11)**.

**Figure 3:**
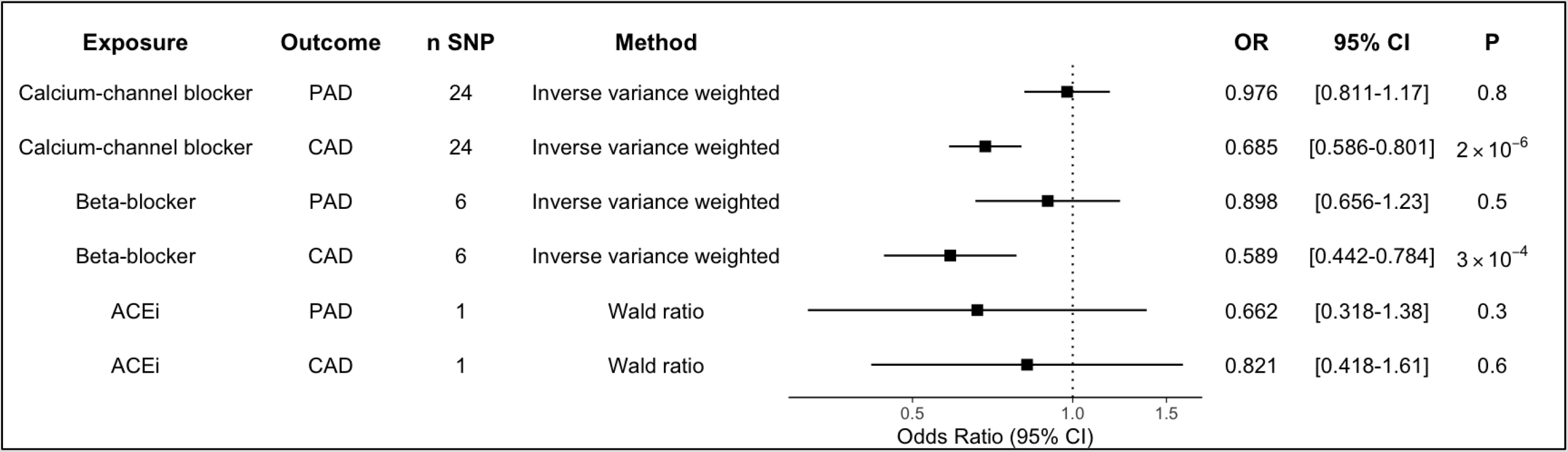
Mendelian Randomization Estimates of the Effect of Systolic Blood Pressure Lowering Through Antihypertensive Medication-associated Genetic Variants. Results scaled to reflect odds of outcome per 10mmHg decrease in blood pressure. n SNP = number of single nucleotide polymorphisms in the exposure instrument; OR = odds ratio; 95% CI = 95% confidence interval; P = p-value SBP = Systolic blood pressure; DBP = Diastolic blood pressure; PP = Pulse pressure; PAD = Peripheral artery disease; CAD = coronary artery disease.

In the second analysis, the effects of both SBP and DBP lowering conferred by genetic variants located within targets of a broader set of antihypertensive medications were considered **(Supplemental Tables 12–13)**. Results for SBP were similar to the first analysis, identifying protective effects of calcium channel blocker associated variants on CAD (OR 0.64 per 10mmHg decrease in SBP; 95% CI 0.50–0.81; p = 2 × 10^−4^), vasodilator associated variants on CAD (OR 0.62 per 10mmHg decrease in SBP; 95% CI 0.40–0.95; p = 0.03), and angiotensin II receptor antagonist associated variants on CAD (OR 0.30 per 10mmHg decrease in SBP; 95% CI 0.097–0.95; p = 0.04). Though not associated with CAD, SBP lowering via beta-blocker associated variants decreased risk of PAD (OR 0.51 10mmHg decrease in SBP; 95% CI 0.27–0.97; p = 0.04). Only results for calcium channel blocker associated variants were robust in weighted-median sensitivity analyses.

For diastolic blood pressure, lowering via calcium channel blocker associated variants decreased risk of CAD (OR 0.52 per 10mmHg decrease in DBP; 95% CI 0.34–0.78; p = 0.002), lowering via angiotensin II receptor antagonist associated variants decreased risk of CAD (OR 0.11 per 10mmHg decrease in DBP; 95% CI 0.014–0.91; p = 0.04), lowering via beta-blocker associated variants decreased risk of CAD (OR 0.43 per 10mmHg decrease in DBP; 95% CI 0.19–1; p = 0.05), and lowering via thiazide diuretic associated variants increased risk of PAD (OR 2.97 per 10mmHg decrease in DBP; 95% CI 1.1–8; p = 0.03). Only results for calcium channel blocker associated variants were robust in weighted-median sensitivity analyses **(Supplemental Table 13)**.

## DISCUSSION

This Mendelian randomization study leveraged natural genetic variation in blood pressure in up to 757,601 individuals to examine the relationship between blood pressure and both PAD and CAD. The principal findings were: 1) Lifetime exposure to elevated SBP, DBP, and PP all increased risk of PAD and CAD; 2) Elevated SBP and DBP more strongly increased risk of CAD compared to PAD; 3) PAD led to a small but significant increase in PP; 4) Based on genetic proxies, the optimal antihypertensive regimens for prevention/treatment of PAD remains unclear. There are several implications from the results of this study.

First, this study supports observational findings that elevated blood pressure is associated with increased risk of PAD. Multiple observational studies have identified elevated SBP and clinical diagnosis of hypertension as strong risk factors for PAD, while the relationship between DBP and PAD has remained less clear (5–11,25–27). Unlike other observational studies, our MR study leveraged genetic variants as instrumental variables for SBP, DBP, and PP. Because genetic variants are randomly inherited by offspring from their parents, mimicking a trial randomizing individuals to a lifetime of increased blood pressure, the Mendelian randomization framework is less susceptible to residual environmental confounding than traditional observational studies (14). The finding of our MR analysis that elevated SBP increases risk of both PAD and CAD is consistent with prior studies. We also find a strong effect of DBP on both PAD and CAD, clarifying discrepant findings in prior observational studies. Overall, the MR findings of our study are consistent with a causal relationship between blood pressure traits and both PAD and CAD.

Next, we found that elevated SBP and DBP each increased risk of CAD more than PAD. These findings are in contrast to a prior observational analysis that found that SBP or DBP had similar effects on CAD and PAD (7). While broad recommendations for lifestyle modification and treatment of ASCVD risk factors are clearly important at both the population level and individual level, understanding the impact of interventions on specific ASCVD outcomes may further inform treatment and prevention guidelines and discussions with patients. Particularly in light of our recent finding that smoking more strongly increases risk of PAD in comparison to CAD or ischemic stroke (28), this study adds further nuance to the relationship between traditional ASCVD risk factors and specific ASCVD outcomes.

Our finding that increased pulse pressure increases PAD risk is consistent with findings from multiple prior observational studies (29–32). Because increased pulse pressure is a marker of increased arterial stiffness and may be caused by PAD, the observational studies investigating the relationship between these traits may have been limited by the possibility of reverse causality. Using bi-directional MR we were able to overcome this limitation, finding elevated PP to be a risk factor for PAD, and PAD to be a risk factor for increased PP. Our multivariable MR analysis of SBP and DBP suggests the effect of elevated DBP may be attenuated after accounting for the genetic effect of SBP. In other words, holding diastolic blood pressure constant, the effect of blood pressure on ASCVD is due to the effects of increased SBP (and indirectly, increased PP). These findings lend further support for the role of increased PP on PAD.

Finally, we used antihypertensive drug MR to estimate the effect of 10mmHg lowering of blood pressure by different classes of medication. In this analysis, we identified a protective effect of calcium-channel blockers on risk of CAD, consistent across all sensitivity analyses. Confidence intervals for other drug classes were wide, and did not exclude meaningful effects, which may reflect the small number of genetic variants included in the genetic instruments for each antihypertensive drug class. More robust genetic instruments may ultimately reveal additional antihypertensive drug classes that robustly lower ASCVD risk. Similarly, because increased SBP and DBP more strongly affected risk of CAD than PAD, the lack of effect of medication-specific SBP or DBP-lowering instruments and PAD is not surprising. While small beneficial genetic effects may compound over a lifetime leading to protection from ASCVD, the effects of antihypertensive medications occur on a much shorter timescale. Our findings do not exclude beneficial effects of potent antihypertensive medications on risk of PAD and CAD, particularly given the strong causal effects of each BP trait and each ASCVD outcome.

The overall findings of our study have implications for PAD prevention and treatment guidelines. The current 2016 American Heart Association/American College of Cardiology (AHA/ACC) and 2017 European Society of Cardiology (ESC) PAD guidelines make strong recommendations for the treatment of hypertension to prevent cardiovascular events.(3,4) The trials cited to support these recommendations focused on cardiovascular events broadly, or differences in safety and efficacy between different antihypertensive classes, rather than PAD-specific outcomes (33–41). A Cochrane Review found poor evidence for the use of antihypertensive medications specifically for PAD, though recognized the large benefit of these medications for prevention of cardiovascular events and mortality more broadly (42). Our MR study provides strong evidence consistent with a casual effect of increased blood pressure on PAD. In the absence of large randomized trials of antihypertensive medications focused on PAD-specific outcomes, these results add support for current guideline recommendations. Further, these results may help calibrate the expected benefit that programs to treat hypertension may have on the global burden of PAD.

This study has several potential limitations. The genetic studies of blood pressure, CAD, and PAD used in our analysis were primarily composed of individuals of European ancestry. Further study of BP and ASCVD genetics in diverse ancestral populations is necessary to improve the generalizability of our findings. Mendelian randomization relies on a number of assumptions in order for causal estimates to be valid (14). While we have employed multiple MR methods and sensitivity analyses to assess for and address potential violations of these assumptions, we cannot completely exclude the possibility of confounding. Future study on the role of hypertension treatment in the prevention and treatment of PAD focused on PAD-specific outcomes is warranted.

Overall, we find strong evidence consistent with a causal effect of blood pressure traits on ASCVD outcomes, with a stronger effect of SBP and DBP on CAD in comparison to PAD. Further study of the differential effects of traditional ASCVD risk factors on specific ASCVD outcomes may help guide prevention and treatment strategies for these common diseases.

## Data Availability

Summary statistics for the blood pressure genome wide association study are publicly available, and may be downloaded from the NHLBI GRASP catalog (https://grasp.nhlbi.nih.gov/FullResults.aspx). MVP PAD genome wide association study summary statistics are available on dbGAP (Accession phs001672.v2.p1). Data on coronary artery disease have been contributed by CARDIoGRAMplusC4D investigators and have been downloaded from www.cardiogramplusc4d.org/data-downloads/.

## FUNDING

This work was supported by US Department of Veterans Affairs grants IK2-CX001780 (Damrauer), and I01-BX003362 (Tsao/Chang). This research is based on data from the MVP, Office of Research and Development, Veterans Health Administration and was supported by award no. MVP000. This publication does not represent the views of the Department of Veterans Affairs or the United States government. This work was also supported by the National Institute of Diabetes and Digestive and Kidney Diseases R01-DK101478 (Voight), and a Linda Pechenik Montague Investigator Award (Voight). The Medical Research Council (MRC) and the University of Bristol support the MRC Integrative Epidemiology Unit [MC_UU_12013/1, MC_UU_12013/9, MC_UU_00011/1]. NMD is supported by a Norwegian Research Council Grant number 295989.

## DATA AVAILABILITY

Summary statistics for the blood pressure genome wide association study are publicly available, and may be downloaded from the NHLBI GRASP catalog

(https://grasp.nhlbi.nih.gov/FullResults.aspx). MVP PAD genome wide association study summary statistics are available on dbGAP (Accession phs001672.v2.p1). Data on coronary artery disease have been contributed by CARDIoGRAMplusC4D investigators and have been downloaded from http://www.cardiogramplusc4d.org/data-downloads/.

---

**Supplemental Figure 1:**
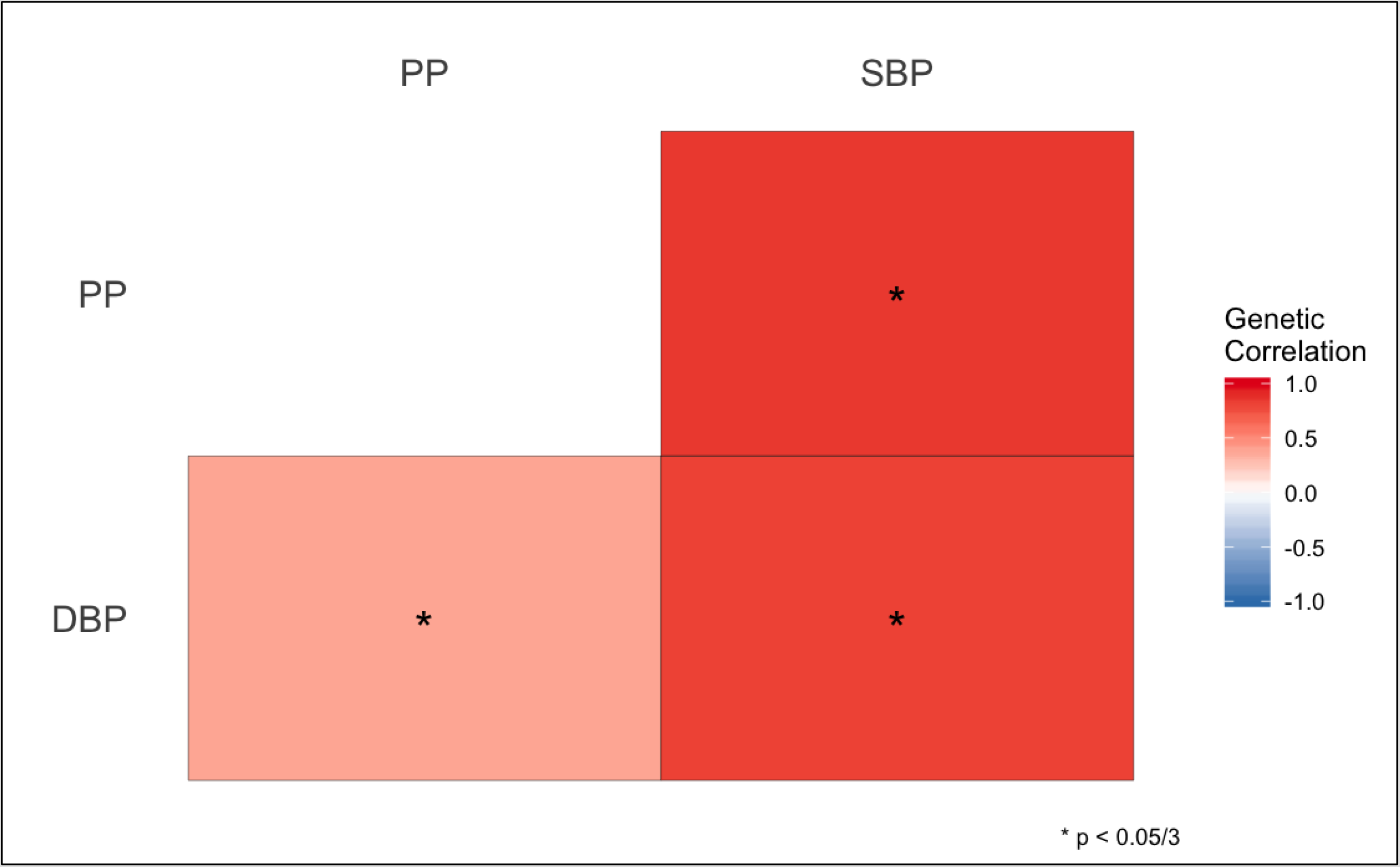
Genetic Correlation Between Blood Pressure Traits

**Supplemental Figure 2:**
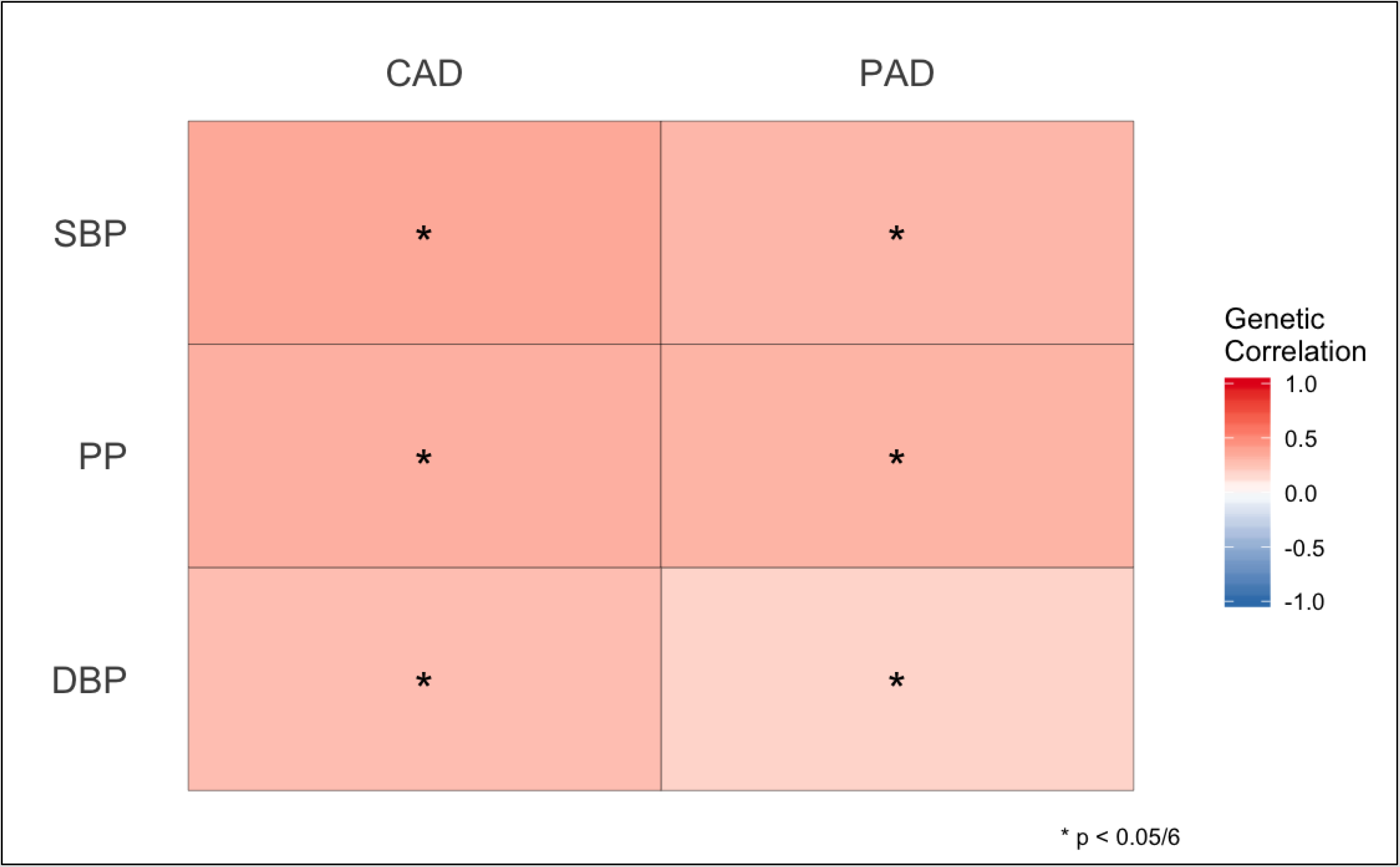
Genetic Correlation Between Blood Pressure and ASCVD Traits

## Notes

### Competing Interest Statement

The authors have declared no competing interest.

### Author Declarations

This study utilized only publicly available, de-identified summary statistics from previously published works, making it exempt from Institutional Review Board review at the University of Pennsylvania.

## REFERENCES

1. James SL, Abate D, Abate KH, Abay SM, Abbafati C, Abbasi N, et al. Global, regional, and national incidence, prevalence, and years lived with disability for 354 Diseases and Injuries for 195 countries and territories, 1990–2017: A systematic analysis for the Global Burden of Disease Study 2017. Lancet. 2018 Nov 10;1789–858.

2. Criqui MH, Aboyans V. Epidemiology of Peripheral Artery Disease. Circ Res. 2015 Apr 24;116(9):1509–26.

3. Gerhard-Herman MD, Gornik HL, Barrett C, Barshes NR, Corriere MA, Drachman DE, et al. 2016 Lower Extremity Peripheral Arterial Disease Guidelines. J Am Coll Cardiol. 2017;69(11):e71–126.

4. Aboyans V, Ricco J-B, Bartelink M-LEL, Björck M, Brodmann M, Cohnert T, et al. 2017 ESC Guidelines on the Diagnosis and Treatment of Peripheral Arterial Diseases, in collaboration with the European Society for Vascular Surgery (ESVS): Document covering atherosclerotic disease of extracranial carotid and vertebral, mesenteric, renal,. Eur Heart J [Internet]. 2017 Aug 26;39(9):763–816. Available from: https://doi.org/10.1093/eurheartj/ehx095

5. Bainton D, Sweetnam P, Baker I, Elwood P. Peripheral vascular disease: consequence for survival and association with risk factors in the Speedwell prospective heart disease study. Heart [Internet]. 1994 Aug 1 [cited 2019 Oct 10];72(2):128–32. Available from: http://heart.bmj.com/cgi/doi/10.1136/hrt.72.2.128

6. Criqui MH, Vargas V, Denenberg JO, Ho E, Allison M, Langer RD, et al. Ethnicity and peripheral arterial disease: The San Diego population study. Circulation. 2005 Oct 25;112(17):2703–7.

7. Gerald F, Housley E, Riemersma RA, Macintyre CCA, Cawood EHH, Prescott RJ, et al. Smoking, lipids, glucose intolerance, and blood pressure as risk factors for peripheral atherosclerosis compared with ischemic heart disease in the Edinburgh artery study. Am J Epidemiol. 1992 Feb 15;135(4):331–40.

8. Hooi JD, Kester ADM, Stoffers HEJH, Overdijk MM, Van Ree JW, Knottnerus JA. Incidence of and risk factors for asymptomatic peripheral arterial occlusive disease: A longitudinal study. Am J Epidemiol. 2001 Apr 1;153(7):666–72.

9. Dagenais GR, Maurice S, Robitaille NM, Gingras S, Lupien PJ. Intermittent claudication in Quebec men from 1974–1986: the Quebec Cardiovascular Study. Clin Invest Med [Internet]. 1991 Apr [cited 2019 Oct 10];14(2):93–100. Available from: http://www.ncbi.nlm.nih.gov/pubmed/2060193

10. Garg PK, Biggs ML, Carnethon M, Ix JH, Criqui MH, Britton KA, et al. Metabolic syndrome and risk of incident peripheral artery disease: The cardiovascular health study. Hypertension. 2014 Feb;63(2):413–9.

11. D.P.J. H, A. B, J.F. F, L. H, L.E. S, P.M. R, et al. Population-Based Study of Incidence, Risk Factors, Outcome, and Prognosis of Ischemic Peripheral Arterial Events: Implications for Prevention. Circulation [Internet]. 2015 [cited 2019 Oct 10];132(19):1805–15. Available from: http://www.embase.com/search/results?subaction=viewrecord&from=export&id=L606470115

12. Klarin D, Lynch J, Aragam K, Chaffin M, Assimes TL, Huang J, et al. Genome-wide association study of peripheral artery disease in the Million Veteran Program. Nat Med [Internet]. 2019 Jul 8 [cited 2019 Jul 28]; Available from: http://www.nature.com/articles/s41591-019-0492-5

13. Nikpay M, Goel A, Won HH, Hall LM, Willenborg C, Kanoni S, et al. A comprehensive 1000 Genomes-based genome-wide association meta-analysis of coronary artery disease. Nat Genet. 2015;47(10):1121–30.

14. Davies NM, Holmes M V, Davey Smith G. Reading Mendelian randomisation studies: A guide, glossary, and checklist for clinicians. BMJ [Internet]. 2018 Jul 12 [cited 2019 Feb 15];362:k601. Available from: http://www.ncbi.nlm.nih.gov/pubmed/30002074

15. Evangelou E, Warren HR, Mosen-Ansorena D, Mifsud B, Pazoki R, Gao H, et al. Genetic analysis of over 1 million people identifies 535 new loci associated with blood pressure traits. Nat Genet. 2018 Oct 1;50(10):1412–25.

16. Bulik-Sullivan BK, Loh P-R, Finucane HK, Ripke S, Yang J, Patterson N, et al. LD Score regression distinguishes confounding from polygenicity in genome-wide association studies. Nat Genet [Internet]. 2015 Mar 2 [cited 2019 Feb 13];47(3):291–5. Available from: http://www.nature.com/articles/ng.3211

17. Bulik-Sullivan B, Finucane HK, Anttila V, Gusev A, Day FR, Loh PR, et al. An atlas of genetic correlations across human diseases and traits. Nat Genet. 2015 Nov 1;47(11):1236–41.

18. Hemani G, Zheng J, Elsworth B, Wade KH, Haberland V, Baird D, et al. The MR-base platform supports systematic causal inference across the human phenome. Elife. 2018;

19. Shim H, Chasman DI, Smith JD, Mora S, Ridker PM, Nickerson DA, et al. A multivariate genome-wide association analysis of 10 LDL subfractions, and their response to statin treatment, in 1868 Caucasians. PLoS One. 2015;

20. Palmer TM, Lawlor DA, Harbord RM, Sheehan NA, Tobias JH, Timpson NJ, et al. Using multiple genetic variants as instrumental variables for modifiable risk factors. Stat Methods Med Res [Internet]. 2012 Jun 7 [cited 2019 Mar 24];21(3):223–42. Available from: http://journals.sagepub.com/doi/10.1177/0962280210394459

21. Bowden J, Davey Smith G, Haycock PC, Burgess S. Consistent Estimation in Mendelian Randomization with Some Invalid Instruments Using a Weighted Median Estimator. Genet Epidemiol [Internet]. 2016 May [cited 2019 Feb 28];40(4):304–14. Available from: http://www.ncbi.nlm.nih.gov/pubmed/27061298

22. Sanderson E, Davey Smith G, Windmeijer F, Bowden J. An examination of multivariable Mendelian randomization in the single-sample and two-sample summary data settings. Int J Epidemiol. 2019 Jun 1;48(3):713–27.

23. Gill D, Georgakis MK, Koskeridis F, Jiang L, Feng Q, Wei W-Q, et al. Use of Genetic Variants Related to Antihypertensive Drugs to Inform on Efficacy and Side Effects. Circulation. 2019 Jul 23;140(4):270–9.

24. Walker VM, Kehoe PG, Martin RM, Davies NM. Repurposing antihypertensive drugs for the prevention of Alzheimer’s disease: a Mendelian randomization study. Int J Epidemiol. 2019 Jul 23;

25. Meijer WT, Grobbee DE, Hunink MGM, Hofman A, Hoes AW. Determinants of peripheral arterial disease in the elderly: The Rotterdam Study. Arch Intern Med. 2000 Oct 23;160(19):2934–8.

26. Newman AB, Siscovick DS, Manolio TA, Polak J, Fried LP, Borhani NO, et al. Ankle-arm index as a marker of atherosclerosis in the cardiovascular health study. Circulation. 1993;88(3):837–45.

27. McGee DL. Update on Some Epidemiologic Features of Intermittent Claudication: The Framingham Study. J Am Geriatr Soc. 1985;33(1):13–8.

28. Levin MG, Klarin D, Assimes TL, Freiberg MS, Ingelsson E, Lynch J, et al. Genetics of Smoking and Risk of Atherosclerotic Cardiovascular Diseases: A Mendelian Randomization Study. medRxiv. 2020 Apr 8;2020.04.07.20053447.

29. Mao Y, Huang Y, Yu H, Xu P, Yu G, Yu J, et al. Incidence of peripheral arterial disease and its association with pulse pressure: A prospective cohort study. Front Endocrinol (Lausanne). 2017 Nov 24;8(NOV).

30. Zhan Y, Yu J, Chen R, Sun Y, Fu Y, Zhang L, et al. Prevalence of low ankle brachial index and its association with pulse pressure in an elderly Chinese population: a cross-sectional study. J Epidemiol [Internet]. 2012 [cited 2019 Oct 10];22(5):454–61. Available from: http://www.ncbi.nlm.nih.gov/pubmed/22813646

31. Korhonen P, Kautiainen H, Aarnio P. Pulse pressure and subclinical peripheral artery disease. J Hum Hypertens [Internet]. 2014 Apr [cited 2019 Oct 10];28(4):242–5. Available from: http://www.ncbi.nlm.nih.gov/pubmed/24132137

32. Kiuchi S, Hisatake S, Watanabe I, Toda M, Kabuki T, Oka T, et al. Pulse Pressure and Upstroke Time Are Useful Parameters for the Diagnosis of Peripheral Artery Disease in Patients With Normal Ankle Brachial Index. Cardiol Res [Internet]. 2016 Oct [cited 2019 Oct 10];7(5):161–6. Available from: http://www.ncbi.nlm.nih.gov/pubmed/28197286

33. Ostergren J, Sleight P, Dagenais G, Danisa K, Bosch J, Qilong Y, et al. Impact of ramipril in patients with evidence of clinical or subclinical peripheral arterial disease. Eur Heart J [Internet]. 2004 Jan [cited 2019 Oct 10];25(1):17–24. Available from: http://www.ncbi.nlm.nih.gov/pubmed/14683738

34. Yusuf S. Effects of an angiotensin-converting-enzyme inhibitor, ramipril, on cardiovascular events in high-risk patients. N Engl J Med. 2000 Jan 20;342(3):145–53.

35. Yusuf S, Teo KK, Pogue J, Dyal L, Copland I, Schumacher H, et al. Telmisartan, ramipril, or both in patients at high risk for vascular events. N Engl J Med. 2008 Apr 10;358(15):1547–59.

36. Bavry AA, Anderson RD, Gong Y, Denardo SJ, Cooper-Dehoff RM, Handberg EM, et al. Outcomes Among hypertensive patients with concomitant peripheral and coronary artery disease: findings from the INternational VErapamil-SR/Trandolapril STudy. Hypertens (Dallas, Tex 1979) [Internet]. 2010 Jan [cited 2019 Oct 10];55(1):48–53. Available from: http://www.ncbi.nlm.nih.gov/pubmed/19996066

37. Zanchetti A, Julius S, Kjeldsen S, McInnes GT, Hua T, Weber M, et al. Outcomes in subgroups of hypertensive patients treated with regimens based on valsartan and amlodipine: An analysis of findings from the VALUE trial. J Hypertens [Internet]. 2006 Nov [cited 2019 Oct 10];24(11):2163–8. Available from: http://www.ncbi.nlm.nih.gov/pubmed/17053536

38. Diehm C, Pittrow D, Lawall H. Effect of nebivolol vs. hydrochlorothiazide on the walking capacity in hypertensive patients with intermittent claudication. J Hypertens [Internet]. 2011 Jul [cited 2019 Oct 10];29(7):1448–56. Available from: http://www.ncbi.nlm.nih.gov/pubmed/21602713

39. Espinola-Klein C, Weisser G, Jagodzinski A, Savvidis S, Warnholtz A, Ostad M-A, et al. β-Blockers in patients with intermittent claudication and arterial hypertension: results from the nebivolol or metoprolol in arterial occlusive disease trial. Hypertens (Dallas, Tex 1979) [Internet]. 2011 Aug [cited 2019 Oct 10];58(2):148–54. Available from: http://www.ncbi.nlm.nih.gov/pubmed/21646599

40. Paravastu SCV, Mendonca DA, Da Silva A. Beta blockers for peripheral arterial disease. Cochrane database Syst Rev [Internet]. 2013 Sep 11 [cited 2019 Oct 10];(9):CD005508. Available from: http://www.ncbi.nlm.nih.gov/pubmed/24027118

41. Group TAO and C for the ACR, Coordinators TAO and, Antihypertensive T, Treatment L. Major Outcomes in High-Risk Hypertensive Patients Randomized to Angiotensin-Converting Enzyme Inhibitor or Calcium Channel Blocker vs Diuretic. JAMA J Am Med Assoc [Internet]. 2002 [cited 2019 Oct 10];288(23):2981–97. Available from: http://www.ncbi.nlm.nih.gov/pubmed/12479763

42. Lane DA, Lip GYH. Treatment of hypertension in peripheral arterial disease. Vol. 2013, Cochrane Database of Systematic Reviews. John Wiley and Sons Ltd; 2013.

